# Structural differences between healthy subjects and patients with schizophrenia and schizoaffective disorder: A graph and control theoretical perspective

**DOI:** 10.1101/2021.02.17.21251624

**Authors:** Cristiana Dimulescu, Serdar Gareayaghi, Fabian Kamp, Sophie Fromm, Klaus Obermayer, Christoph Metzner

## Abstract

The coordinated, dynamical interactions of large-scale networks give rise to cognitive function. Recent advances in network neuroscience have suggested that the anatomical organization of such networks puts a fundamental constraint on the dynamical landscape of the brain. Consequently, changes in large-scale brain activity have been hypothesized to underlie many neurological and psychiatric disorders. Specifically, evidence has emerged that large-scale dysconnectivity might play a crucial role in the pathophysiology of schizophrenia. To investigate potential differences in graph and control theoretic measures between patients with schizophrenia (SCZ), patients with schizoaffective disorder (SCZaff) and matched healthy controls (HC), we use structural MRI data. More specifically, we first calculate seven graph measures of integration, segregation, centrality and resilience and test for group differences. Second, we extend our analysis beyond these traditional measures and employ a simplified noise-free linear discrete-time and time-invariant network model to calculate two complementary measures of controllability. Average controllability identifies brain areas that can guide brain activity into different, easily reachable states with little input energy. Modal controllability on the other hand, characterizes regions that can push the brain into difficult-to-reach states, i.e. states that require substantial input energy. We identified differences in standard network and controllability measures for both patient groups compared to HCs. Specifically, we found a strong reduction of betweenness centrality for both patient groups and a strong reduction in average controllability for the SCZ group again in comparison to the HC group. Our findings of network level deficits might help to explain the many cognitive deficits associated with these disorders.

## 1 Introduction

Ample evidence has emerged that dysconnectivity, i.e. network-level abnormalities in the connectivity between brain regions, might play a central role in the pathophysiology of schizophrenia [5, 9]. While traditionally analysis of structural connectivity has looked at gray and white matter volume and tissue anisotropy, over the last decades, network neuroscience, interpreting the brain as a complex network of interconnected regions, has emerged and very successfully applied graph and network science to understand structure-function relationships in the brain [20]. A graph-theoretical analysis of brain networks naturally lends itself to address the dysconnectivity hypothesis in schizophrenia. Overall, studies applying network science techniques have identified disturbances regarding integration and segregation properties in structural and functional networks in schizophrenia [2, 3, 16]. Moving beyond a mere graph theoretical perspective Gu et al. [11], employed a control-theoretic framework to gain a deeper understanding of the dynamic interactions between large-scale brain networks and their relation to cognitive abilities. Their results imply that densely connected areas (e.g. in the default mode network) enable the movement of the brain to many easily reachable states, whereas weakly connected regions (e.g. in the cognitive control systems) help the brain to transition to states that are hard to reach. The strong and widespread deficits in cognitive processes in patients with schizophrenia suggest a global dysfunction of large-scale brain dynamics. Therefore, we hypothesized that patients might display disturbances in measures of network controllability.

To address this hypothesis, we built structural brain networks from DTI and T1w MRI using a freely available data set from the Center of Biomedical Research Excellence (COBRE) data repository of patients with schizophrenia and schizoaffective disorder and subsequently performed a graph- and control-theoretic analysis. We investigated graph measures of integration, segregation, centrality and resilience together with two measures of controllability, average and modal controllability. We found a strong difference between healthy controls and patients for betweenness centrality and, importantly, strong differences in controllability between the groups.

## 2 Methods

### Data Set

We obtained a final sample of 43 healthy control subjects, 43 patients with schizophrenia and 9 patients with schizoaffective disorder from the SchizConnect database (*http://schizconnect.org*). Patients received antipsychotic medication. Symptom severity in patients was assessed using the Positive and Negative Syndrome Scale (PANSS) [14]. Written informed consent was obtained from all participants, and the study was approved by the regional ethics committee. The groups did not differ significantly in terms of age and gender, and the patient groups also did not differ significantly in terms of symptoms as measured with PANSS (see Table 1).

**Table 1:** Demographics and clinical characteristics. Data are shown as mean(standard deviation). Age differences between groups were compared using an independent samples t-test and differences in gender distribution using a chi-square test.

| | HC | SCZ | SCZaff | Statistics, $p$ value |
| --- | --- | --- | --- | --- |
| Group size | 43 | 43 | 9 |  |
| Age | 36.70(11.04) | 38.72(14.02) | 39.00(12.60) | HC/SCZ: $t=-0.73$, $p=0.46$ |
| (years) | | | | HC/SCZaff: $t=-0.54$, $p=0.59$ |
| Gender | 11/32 | 12/31 | 2/7 | HC/SCZ: $\chi^2=0.06$, $p=0.81$ |
| (female/male) | | | | HC/SCZaff: $\chi^2=0.04$, $p=0.83$ |
| PANSS positive | - | 14.34(4.94) | 15.78(3.12) | SCZ/SCZaff: $t=-0.75$, $p=0.45$ |
| PANSS negative | - | 14.45(4.90) | 16.33(4.45) | SCZ/SCZaff: $t=-1.04$, $p=0.30$ |
| PANSS general | - | 28.47(8.16) | 33.78(8.98) | SCZ/SCZaff: $t=-1.65$, $p=0.10$ |
| PANSS total | - | 57.53(13.44) | 65.89(14.93) | SCZ/SCZaff: $t=-1.63$, $p=0.11$ |

Data collection was performed using a Siemens Magnetom Trio 3T MR scanner. Structural images (high resolution T1-weighted) were acquired using a five-echo MPRAGE sequence with the following parameters: repetition time (TR) = 2530 ms; echo time (TE) = 1.64, 3.5, 5.36, 7.22, 9.08 ms; inversion time (TI) = 1200 ms; flip angle (FA) = 7°; field of view (FOV) = 256 mm × 256 mm; matrix = 256 × 256; slice thickness = 1 mm; 192 sagittal slices. Diffusion tensor imaging (DTI) data were acquired using a single-shot EPI sequence with TR/TE = 9000/84 ms; FA = 90°; FOV =256 mm × 256 mm; matrix = 128 × 128; slice thickness = 2 mm without gap; 72 axial slices; 30 non-collinear diffusion gradients (b = 800 s/mm2) and 5 non-diffusion-weighted images(b = 0 s/mm2) equally interspersed between the 30 gradient directions. For more information see also [6].

### Data Preprocessing and Tractography

We conducted preprocessing of anatomical and diffusion images using a semi-automatic pipeline implemented in the FSL toolbox (www.fmrib.ox.ac.uk/fsl, FMRIB, Oxford). For the anatomical images, preprocessing included removal of non-brain tissue and brain extraction using the brain extraction toolbox (BET) implemented in FSL, as well as the generation of a brain mask. After extraction, images were checked for quality and subsequently, 94 cortical and subcortical regions were defined according to the automated anatomical atlas (AAL2) described in [18].

For diffusion data, we performed brain extraction on the b0 images using BET. After correcting the data for head movement and eddy current distortions, we fitted a probabilistic diffusion model to the data by using the Bayesian Estimation of Diffusion Parameters Obtained using Sampling Techniques (BEDPOSTX) FSL toolbox. Finally, we linearly registered each b0 image to the corresponding subject’s anatomical T1 image, transformed the high-resolution anatomical mask volumes containing the cortical parcellations to the subject’s diffusion space, and ran probabilistic tractography with 5000 samples per voxel using FSL’s PROBTRACKX algorithm [4].

### Network Construction

We defined the weight of a connection between two regions *i* and *j* to be the inverse of the normalized number of fibres *nf*_*ij*_ determined through the probabilistic tracking:

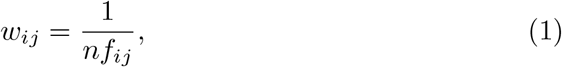

if *nf*_*ij*_ > *T*, with a certain threshold *T*. If not mentioned otherwise, we used a threshold of *T* = 0.Using this definition, the larger the weight between two regions the weaker the connectivity is between these regions, or in other words, the higher cost is to go from one node to the other. This approach is similar to previously used definitions of weighted structural networks [23, 1, 10].

### Network Measures

We followed Rubinov and Sporns [19] and defined the network measures as follows. Let *W* = *w*_*ij*_ be the weighted matrix describing the pair-wise connectivity between regions constructed from the DTI scans. Furthermore, let *N* be the set of nodes in the network with *n* := |*N* | = 94. Then we can define some basic measures describing a network. First, the **weighted degree** of node *i* can be defined as

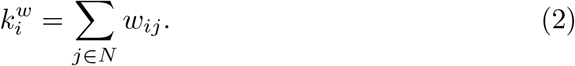

The **shortest weighted path length** between two nodes *i* and *j* is given by

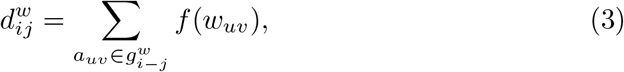

where f is the inverse of the weight and 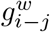 is the shortest weighted path between *i* and *j*. Lastly, **weighted geometric mean of triangles** around a node *i* can be calculated as

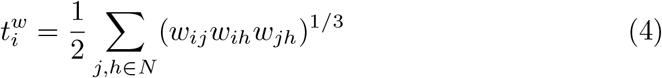

With these basic measures we can now define the network measures of interest.

First, we define two measures of integration: **characteristic path length** and **global efficiency**. The **weighted characteristic path length** describes the average shortest path length between two nodes in the network and is defined as

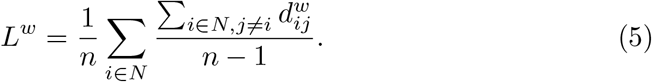

The **global efficiency** is defined as

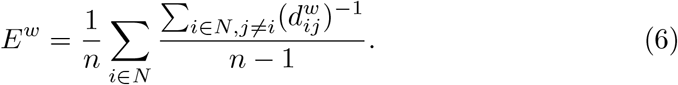

Next, we define two measures of segregation: the **weighted clustering coefficient** and the **weighted transitivity**. The **weighted clustering coefficient** can be calculated as

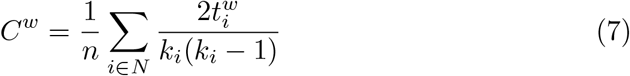

and the **weighted transitivity** is given by

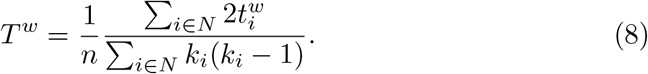

We then introduce two measures of centrality: **average closeness centrality** and **average betweenness centrality**. We define **average closeness centrality** as the average of nodal **closeness centrality** over all nodes, where **closeness centrality** of a node *i* is given by

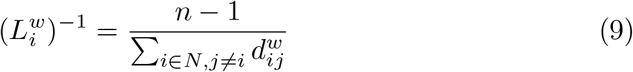

and, similarly, **average betweenness centrality** as the average of nodal **betweenness centrality** over all nodes, where **betweenness centrality** of a node *i* calculated as

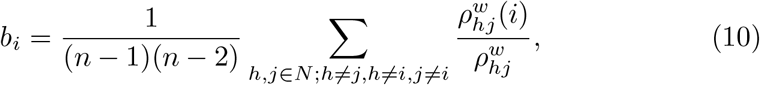

where 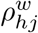 is the number of shortest weighted paths between *h* and *j*, and 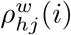 is the number of shortest weighted paths between *h* and *j* that pass through *i*.

Lastly, we also defined a measure of network resilience:**weighted assortativity. Weighted assortativity** can be defined as

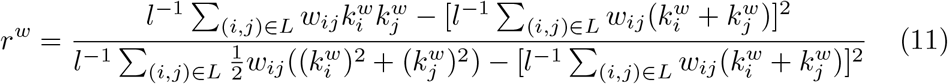

### Controllability Measures

Here we utilized the control theoretic notion of controllability which gives theoretical insight into the effect local changes of dynamics might have. We chose two different measures based on a linear control setting: *average controllability* and *modal controllability*. **Average controllability** is given by the average input energy to the control nodes over all possible target states. Typically, nodes with high average controllability can control networks dynamics over nearby target states in an energy efficient way. On the other hand, **modal controllability** describes how well a node can control all network modes. High modal controllability means that a node is able to reach all modes of a network and, thus, can force the dynamics into hard-to-reach target states.

To define the controllability measures, we assume a simplified noise-free linear discrete-time and time-invariant network model of the form

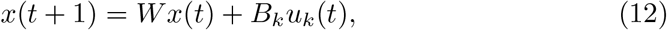

where *x* is the state of the network, *W* is again the weighted adjacency matrix of the network, the input matrix *B*_*k*_ specifies the control nodes and *u*_*k*_ defines the control strategy over time. In our setting, we always choose a single control node *i*, which means that *B*_*i*_ is simply a unit vector with entry at row *i*. This allows us to define the so-called **controllability Gramian** as

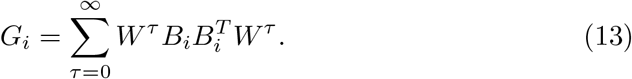

We then define **average controllability** of node *i* simply as the trace *Tr*(*G*_*i*_) of the controllability Gramian (see [17, 11]). The **average controllability** of the network is then defined as the mean of the average controllability over all nodes in the network. **Modal controllability** of node *i* will be defined as

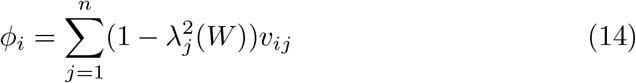

where *V* = (*υ*_*ij*_) is the matrix of eigenvectors from the weighted adjacency matrix *W* and λ_*j*_(*W*) are the eigenvalues of *W*.

## 3 Results

### 3.1 Graph Measures

We first compared seven standard graph measures in the two patient groups against the shared control of healthy subjects. The graph measures were: characteristic path length and global efficiency (two measures of integration), cluster coefficient and transitivity (two measures of segregation), betweenness and closeness centrality (two measures of centrality), and assortativity (a measure of resilience). Patients with schizophrenia showed a strong reduction of betweenness centrality compared to healthy controls (*g* = *−*0.644, *p <* 0.05 Bonferroni corrected; see also Figure 1 and Table 2). In our exploratory analysis of the small schizoaffective group, patients showed a similar reduction in betweenness centrality than the SCZ group (*g* = *−*1.080, *p <* 0.05 Bonferroni corrected; see also Figure 1 and Table 3). Interestingly, they also displayed an increase in characteristic path length, which, however, did not survive correction for multiple comparisons (*g* = 0.766, *p* = 0.266 Bonferroni corrected; see also Figure 1 and Table 3) Furthermore, note the small sample size of the schizoaffective group.

**Table 2:** Permutation test statistics for the SCZ group(sample size *n* = 43). Comparison against the HC group (sample size *n* = 43) (see [13]).

| | Hedge's $g$ | 95% CI interval | permutation p-value<br>(uncorrected) |
| --- | --- | --- | --- |
| Characteristic path length | 0.385 | $[-0.056, 0.814]$ | $p = 0.079$ |
| Cluster coefficient | -0.406 | $[-0.715, -0.017]$ | $p = 0.056$ |
| Betweenness centrality | -0.644 | $[-1.070, -0.204]$ | $p = 0.004$ |
| Closeness centrality | -0.384 | $[-0.808, 0.059]$ | $p = 0.078$ |
| Transitivity | -0.340 | $[-0.753, -0.091]$ | $p = 0.110$ |
| Global efficiency | -0.381 | $[-0.802, 0.058]$ | $p = 0.081$ |
| Assortativity | -0.198 | $[-0.605, 0.226]$ | $p = 0.363$ |

**Table 3:** Permutation test statistics for the SCZaff group (sample size *n* = 9). Comparison against the HC group (sample size *n* = 43) (see [13]).

| | Hedge's $g$ | 95% CI interval | permutation p-value<br>(uncorrected) |
| --- | --- | --- | --- |
| Characteristic path length | 0.766 | $[-0.206, 1.480]$ | $p = 0.038$ |
| Cluster coefficient | -0.371 | $[-0.770, 0.473]$ | $p = 0.293$ |
| Betweenness centrality | -1.080 | $[-1.810, -0.049]$ | $p = 0.005$ |
| Closeness centrality | -0.626 | $[-1.230, 0.429]$ | $p = 0.090$ |
| Transitivity | -0.505 | $[-1.190, 0.678]$ | $p = 0.173$ |
| Global efficiency | -0.615 | $[-1.230, 0.462]$ | $p = 0.093$ |
| Assortativity | 0.065 | $[-0.461, 0.642]$ | $p = 0.855$ |

**Figure 1:**
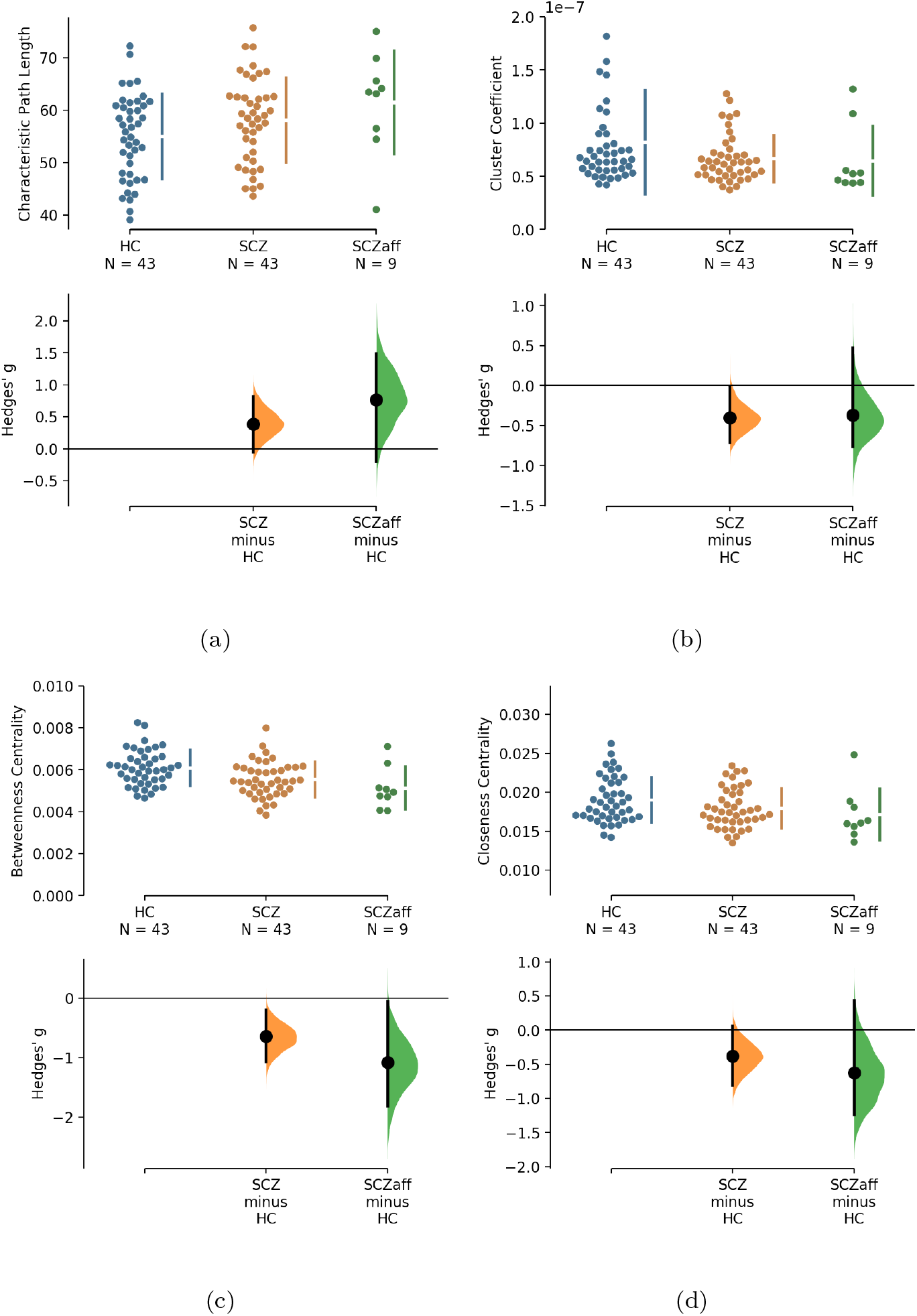
Hedge’s g for the comparisons of various graph measures for the SCZ and SCZaff groups against the HC group are shown in the above Cumming estimation plot. The raw data is plotted on the upper axes. On the lower axes, mean differences are plotted as bootstrap sampling distributions. Each mean difference is depicted as a dot. Each 95% confidence interval is indicated by the ends of the vertical error bars (see [13]).

Next, we explored potential associations between graph measures and symptom scores, i.e. PANSS total, PANSS positive, PANSS negative, and PANSS general scores. We found a moderate positive correlation between assortativity and PANSS total score in the SCZ group (*r* = 0.346, *p* = 0.023; see also Supplementary Figure 5) and a strong negative correlation between assortativity and PANSS positive score in the SCZaff group (*r* = *−*0.846, *p* = 0.004; see Figure 6). Additionally, we found a strong positive correlation between cluster coefficient and PANSS general score (*r* = 0.693, *p* = 0.038; see Supplementary Figure 6) and a moderate negative correlation between the cluster coefficient and the PANSS positive score for the SCZaff group (*r* = *−*0.304, *p* = 0.047; see Supplementary Figure 6). However, none of them survived correction for multiple comparisons. We found no other significant correlations between graph measures and PANSS total scores neither for the SCZ nor for the SCZaff group (see Supplementary Figures 5 and 6).

**Figure 2:**
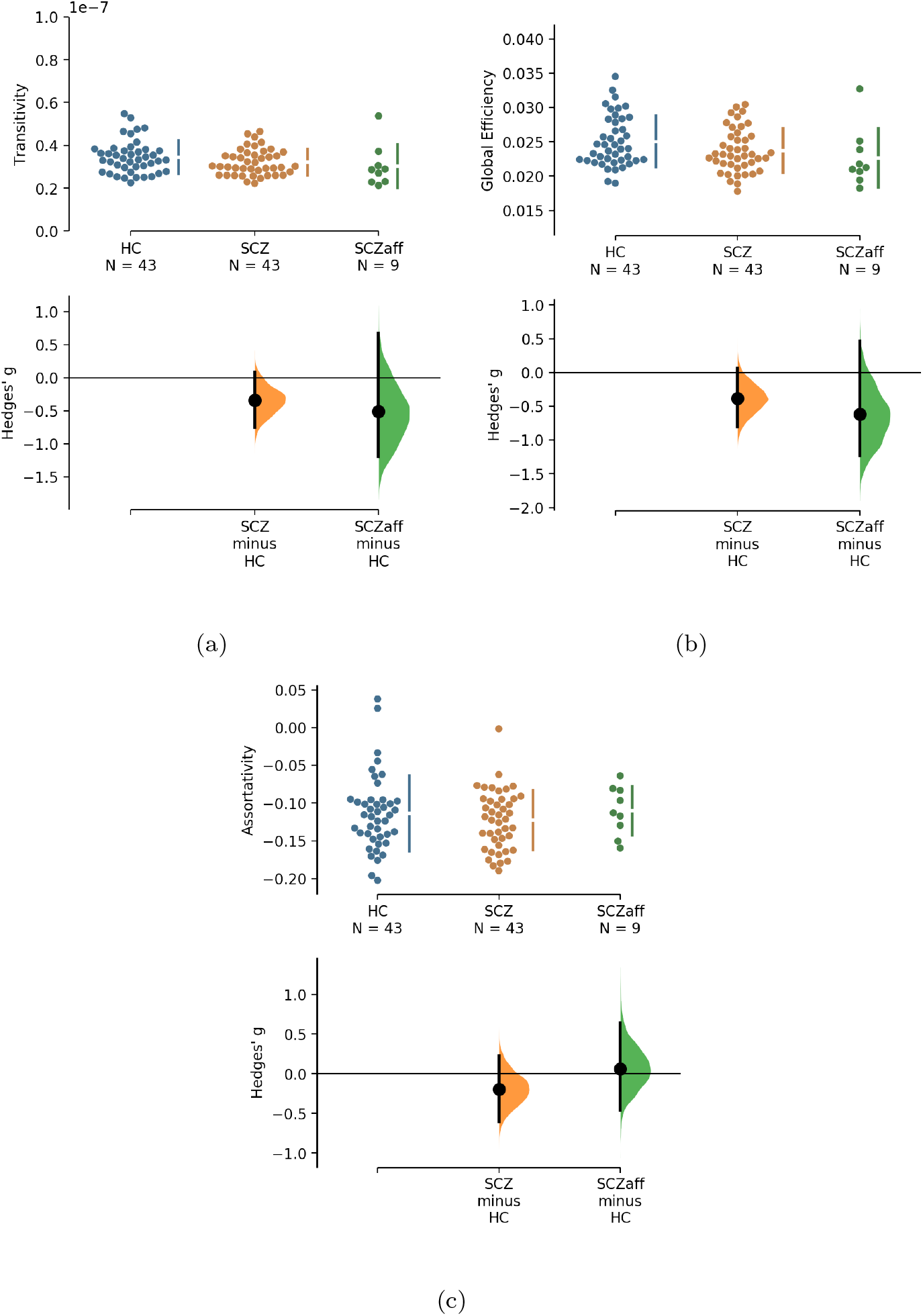
Hedge’s g for the comparisons of various graph measures for the SCZ and SCZaff groups against the HC group are shown in the above Cumming estimation plot. The raw data is plotted on the upper axes. On the lower axes,mean differences are plotted as bootstrap sampling distributions. Each mean difference is depicted as a dot. Each 95% confidence interval is indicated by the ends of the vertical error bars (see [13]).

**Figure 3:**
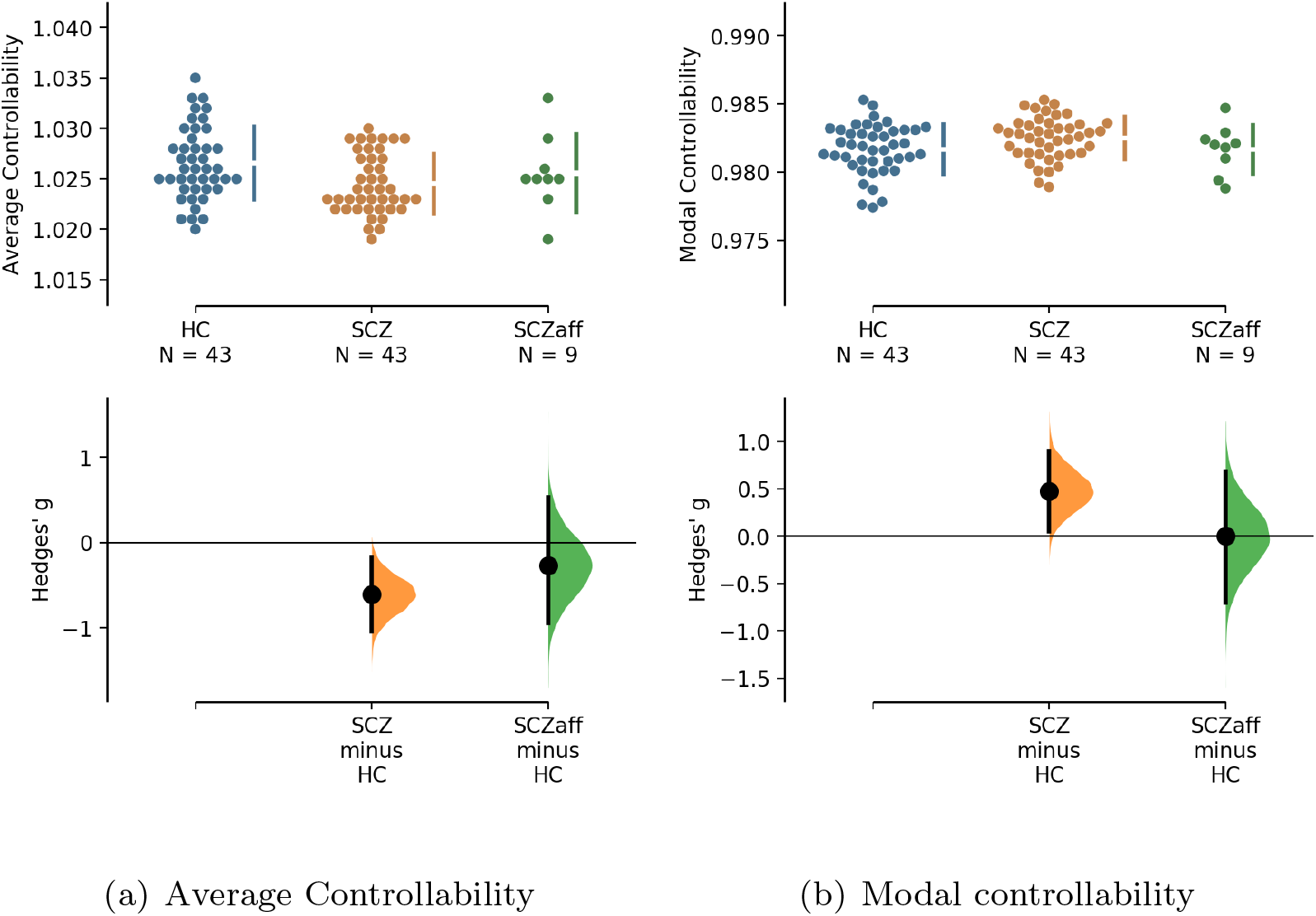
Hedge’s *g* for the comparisons of (a) average and (b) modal controllability of unweighted graphs of the SCZ and SCZaff groups against the HC group are shown in the above Cumming estimation plot. The raw data is plotted on the upper axes. On the lower axes, Hedge’s *g* is plotted as bootstrap sampling distributions. Each *g* is depicted as a dot. Each 95% confidence interval is indicated by the ends of the vertical error bars (see [13]).

**Figure 4:**
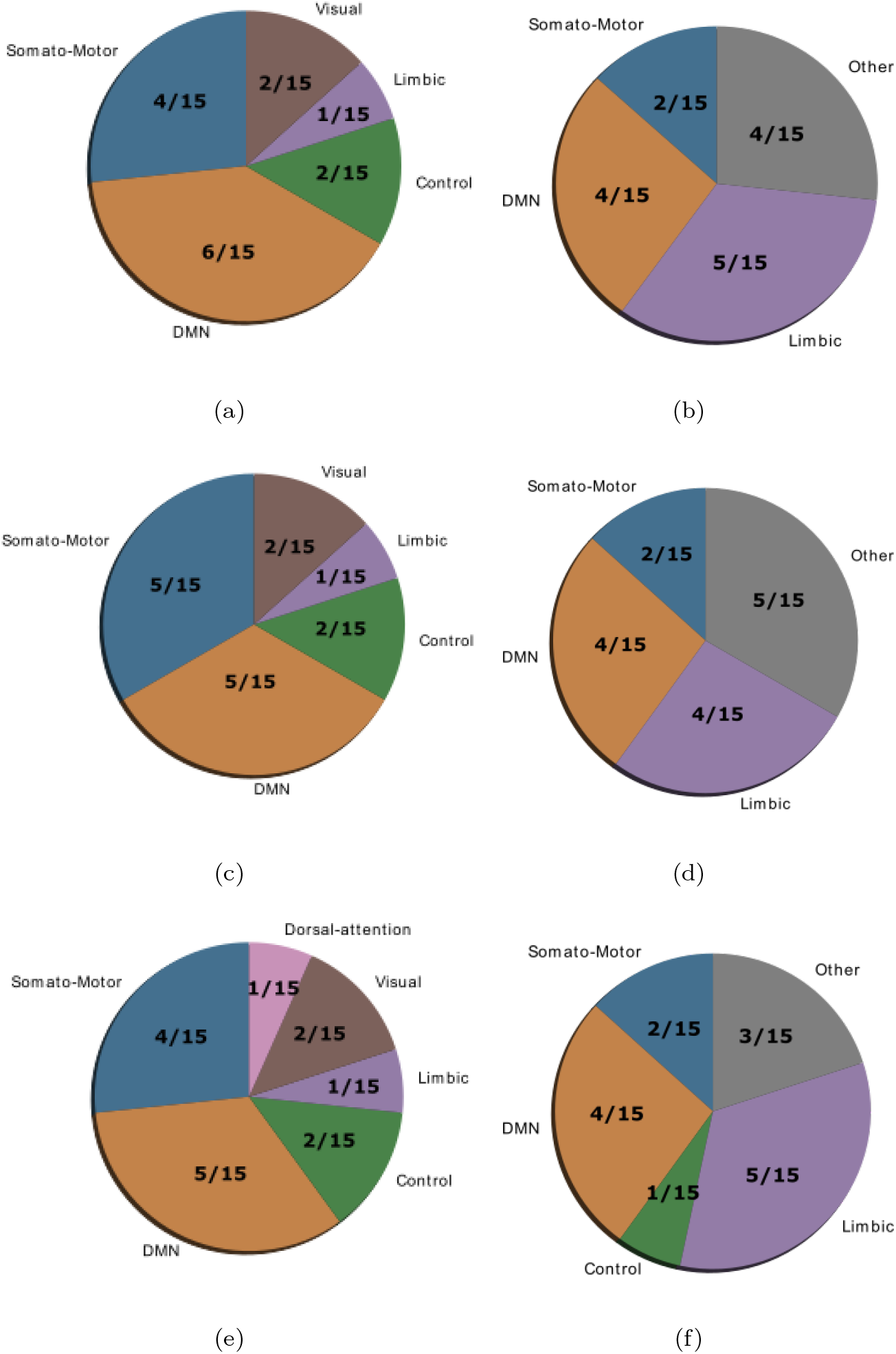
Location of cognitive control hubs for average controllability across large-scale cognitive systems in the HC group (average controllability (a) and modal controllability (b)), the SCZ group (average controllability (c) and modal controllability (d)) and the SCZaff group (average controllability (e) and modal controllability (f)). We chose the top 15 regions with highest average controllability (averaged over all subjects).

### 3.2 Controllability

Going beyond traditional network measures, we also explored two control theoretic measures: average and modal controllability. Network control theory has been increasingly utilized in network neuroscience and offers a mechanistic framework to understand the effects of local changes in dynamics on the global brain state [17]. We specifically chose average and modal controllability because they offer information on two different aspects of controllability: while nodes with a high average controllability have the ability to convey large changes in network dynamics by moving the global system into many easily reachable states, nodes with high modal controllability can move the system to states that are difficult to reach [17].

For the SCZ group we found a strong decrease in average controllability (*g* = *−*0.606, *p <* 0.05 Bonferroni corrected; see also Figure 3 and Table 4). Additionally, we also found a moderate increase of modal controllability in the SCZ group, which, however, did not survive correction for multiple comparisons (*g* = 0.476, *p* = 0.056 Bonferroni corrected; see also Figure 3 and Table 4). We found no differences for the SCZaff group compared to the healthy controls.

**Table 4:** Permutation test statistics for the SCZaff group (sample size *n* = 9). Comparison against the HC group (sample size *n* = 43) (see [13]).

| | Hedge's $g$ | 95% CI | permutation<br>p-value (uncorr.) |
| --- | --- | --- | --- |
| Average controllability<br>(SCZ) | -0.606 | $[-1.040, -0.171]$ | $p = 0.006$ |
| Modal controllability<br>(SCZ) | 0.476 | $[0.047, 0.897]$ | $p = 0.028$ |
| Average controllability<br>(SCZaff) | -0.265 | $[-0.944, 0.537]$ | $p = 0.467$ |
| Modal controllability<br>(SCZaff) | 0.001 | $[-0.701, 0.686]$ | $p = 0.994$ |

**Table 5:** Permutation test statistics for a comparison of the top 15 regions with highest average and modal controllability per group (SCZ and SCZaff), respectively, against the HC group (for details on the permutation testing, see [13]).

| | Hedge's $g$ | 95% CI interval | permutation p-value |
| --- | --- | --- | --- |
| Average controllability (SCZ) | -0.302 | $[-1.030, 0.455]$ | $p = 0.4$ |
| Modal controllability (SCZ) | 0.111 | $[-0.580, 0.858]$ | $p = 0.741$ |
| Average controllability (SCZaff) | -0.054 | $[-0.824, 0.677]$ | $p = 0.877$ |
| Modal controllability (SCZaff) | 0.21 | $[-0.487, 0.922]$ | $p = 0.555$ |

**Table 6:** Assignment of AAL2 regions to one of 8 cognitive systems: Somatomotor, Default Mode (DMN), Control, Visual, Dorsal Attention, Ventral Attention, Limbic, and Other.

| Region | Cognitive System |
| --- | --- |
| Precentral_L | Somatomotor |
| Precentral_R | Somatomotor |
| Frontal_Sup_2_L | Default Mode Network |
| Frontal_Sup_2_R | Default Mode Network |
| Frontal_Mid_2_L | Control |
| Frontal_Mid_2_R | Control |
| Frontal_Inf_Oper_L | Ventral Attention |
| Frontal_Inf_Oper_R | Control |
| Frontal_Inf_Tri_L | Control |
| Frontal_Inf_Tri_R | Control |
| Frontal_Inf_Orb_2_L | Default Mode Network |
| Frontal_Inf_Orb_2_R | Default Mode Network |
| Rolandic_Oper_L | Somatomotor |
| Rolandic_Oper_R | Somatomotor |
| Supp_Motor_Area_L | Somatomotor |
| Supp_Motor_Area_R | Somatomotor |
| Olfactory_L | Limbic |
| Olfactory_R | Limbic |
| Frontal_Sup_Medial_L | Default Mode Network |
| Frontal_Sup_Medial_R | Default Mode Network |
| Frontal_Med_Orb_L | Default Mode Network |
| Frontal_Med_Orb_R | Default Mode Network |

Lastly, we found no correlations between controllability measures and symptom scores, neither for the PANSS total score nor for the positive or negative subscores, in either of the patient groups (see Supplementary Figures 13, 14, 15, and 16).

#### Control Roles of Cognitive Systems

Gu et al. [11] demonstrated that control regions are differentially distributed across cognitive systems depending on whether they show high average or high modal controllability. Therefore, we next asked whether this differential distribution would still be valid in both patient groups or not. To test this, we averaged controllability measures of all 94 regions of the AAL2 atlas (for both average and modal controllability) across all subjects of a group and then extracted the 15 regions with the highest measures. These regions were then assigned one of the following 8 large-scale cognitive systems based on the 7 functional cortical networks from Yeo et al. [25] but extended to also include subcortical structures: Somatomotor, Default Mode (DMN), Control, Visual, Dorsal Attention, Ventral Attention, Limbic, and Other (subcortical regions that could not be assigned to any system; see Supplementary Table 6 for the assignments and [11] for more details on the approach). We found that for average controllability most regions belong to the DMN followed by the somatomotor network in all three groups (Figure 4), consistent with [11]. For modal controllability we found that in all three groups most regions belong to the limbic category (Figure 4). Interestingly, again for all three groups, DMN network regions also made up a large share of the top modal controllability regions. Importantly, the DMN regions with high modal controllability here where distinct from the DMN regions with high average controllability, highlighting the importance and uniqueness of the DMN. Nevertheless, the main finding here is that there are no substantial changes in regions with high average/modal controllability between control and patient groups. Furthermore, there were no strong differences between the mean controllability of the top 15 regions of the different groups, neither for average nor for modal controllability (Supplementary Figure 17 and Table 5).

## 4 Discussion

The brain can be viewed as an interconnected network in which the dynamical transitions between cognitive states give rise to complex behaviours. Psychiatric disorders have been associated with disturbances in these dynamical interactions between brain regions. Schizophrenia for example has been characterized as a disorder of dysconnection [9], where an overall reduction in connectivity leads to disruptions of normal cognitive functions. In this study, we apply techniques from network theory, i.e. standard graph measures, together with novel methods from control theory, i.e. controllability measures, to characterize the degree of dysconnection in structural brain networks derived from white matter fibre connectivity in patients with schizophrenia and a small sample of patients with schizoaffective disorder.

The current study yielded three main findings. First, we found a reduction in betweenness centrality in both patients with schizophrenia and schizoaffective disorder. Since nodes with high betweenness centrality, which quantifies the number of times a node acts as a bridge along shortest paths between pairs of nodes, reflect nodes which can potentially exert strong control over the information flow in the network, the reduction in the patient groups suggests a less controlled flow of information.

Second, structural brain networks from patients with schizophrenia display a strongly reduced average controllability. This finding, which ties nicely with the reduced betweenness centrality, implies that the dysconnectivity in patients with schizophrenia results in a reduced capacity of individual nodes to steer the network activity from one brain state to another. This finding also fits well with the observation that random state switching is more frequent in schizophrenic models, since the lower average controllability implies a reduced capacity of keeping the system in a desired state, which makes random transitions more likely.

Last, the distribution of large-scale functional cognitive systems across nodes with high average or modal controllability is not altered in patient groups, suggesting a global reduction of controllability that does not affect a specific cognitive system. This is not surprising, given that patients with schizophrenia suffer from an array of cognitive symptoms.

Several studies have analysed differences in the structural connectome of patients with schizophrenia using network measures. Overall, these studies report an altered connectome with altered integration and centrality. Filipp et al. [8] found that patient networks were characterized by longer communication pathways and fewer central hubs compared to healthy controls. A reduction in hub structure, especially the hub hierarchy, was also found in a study by Bassett et al. [2]. Furthermore, altered path length between brain nodes and reduced efficiency have consistently been found in schizophrenia [2, 22, 21, 23], with frontal and parietal lobes being most effected. Frontal and parietal lobes were also the areas with the highest reduction of connectivity strength [26]. Overall, our findings are consistent with these previous studies. The reduced betweenness centrality we see would imply a reduction of centrality hubs and their hierarchy, as found by Filippi et al. and Bassett et al. [8, 2] and distort normal communication pathways. Furthermore, the strong decrease of average controllability in schizophrenia patients would be expected in a network where the central hub structure is disturbed. Studies on patients with schizoaffective disorder are rare, however, the affective symptom components in bipolar disorder and schizoaffective disorder are relatively similar. Therefore it is interesting that Collin et al. [7] found the central hub structure to be intact in patients with bipolar disorder. A finding which might explain the absence of differences in average controllability between our schizoaffective patient group and healthy controls, because of the link between central structural hubs, i.e. influential high-degree nodes, and their ability to easily push the brain network into different brain states reflected in their average controllability. However, this absence might be a consequence of the patient sample consisting of stably medicated subjects, since Wang et al. [24] report a disruption of the rich-club hub system in bipolar disorder. Antipsychotic medication has been shown to alter connectivity in SCZ depending on the type of antipsychotic [15]. Furthermore, mood stabilizers tended to renormalize connectivity, while antipsychotic medication did not in patients with bipolar disorder [12]. Therefore, differences in medication in our patient samples might explain the differences with respect to average controllability and central hub structure.

Notably, the current study has its limitations. First, by restricting ourselves to the anatomical parcellation given by the AAL2 atlas, we commit to a specific mapping between MR voxels and brain regions. Importantly, in terms of parcellation schemes, there are a variety of options and no standard has been established so far. Furthermore, taking into account the fact that these schemes differ not only with regard to the number of regions in which they divide the brain, but also with regard to other key aspects, such as the exact location of the inter-regional borders or the method through which they were generated, it is possible that the specific choice of the parcellation scheme has an impact on subsequent graph theoretical findings. Therefore, a reproduction of our findings with other parcellation schemes seems warranted. Second, the patient sample considered in this study consists of chronic, stably medicated subjects and therefore, the findings can only be considered representative of this particular subpopulation of patients. It would indeed be very interesting track changes in network and controllability measures in other patient subpopulations such as first-episode psychosis subjects or throughout the course of the disorders in a longitudinal study design. Furthermore, the presented analysis cannot address whether our findings reflect a primary change due to the disorder or whether they are indeed caused by the antipsychotic medication. Last, the sample size of the current study is not very large and therefore, the results should be confirmed in a replication study with a larger patient group. Importantly, the results from the patients with schizoaffective disorder should be treated with caution because of the very small number of subjects.

This study is, to our best knowledge, the first study investigating controllability measures in patients with schizophrenia and schizoaffective disorder and it corroborates previous findings of altered structural connectivity and suggests that a control theoretic approach could be useful in pathological research.

## Data Availability

Data was obtained through the SchizConnect public database.

http://schizconnect.org/

## Acknowledgements

Data used in preparation of this article were obtained from the SchizConnect database (http://schizconnect.org) As such, the investigators within SchizConnect contributed to the design and implementation of SchizConnect and/or provided data but did not participate in analysis or writing of this report. Data collection and sharing for this project was funded by NIMH cooperative agreement 1U01 MH097435. Data was downloaded from the COllaborative Informatics and Neuroimaging Suite Data Exchange tool (COINS; http://coins.mrn.org/dx) and data collection was performed at the Mind Research Network, and funded by a Center of Biomedical Research Excellence (COBRE) grant 5P20RR021938/P20GM103472 from the NIH to Dr. Vince Calhoun.

## Supplementary Material

### Graph Measures

**Figure 5:**
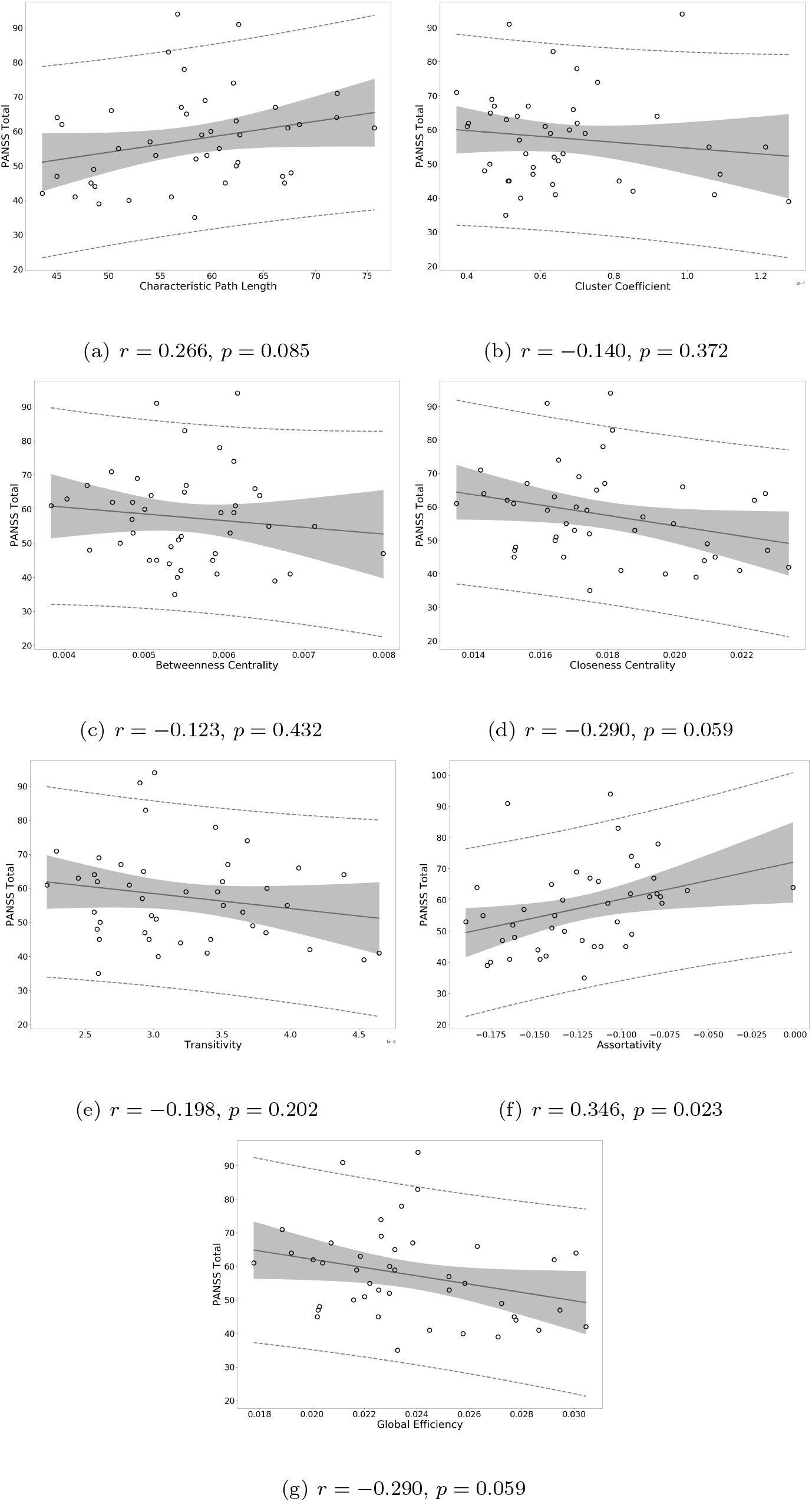
Correlation of graph measures of the SCZ group with PANSS total scores

**Figure 6:**
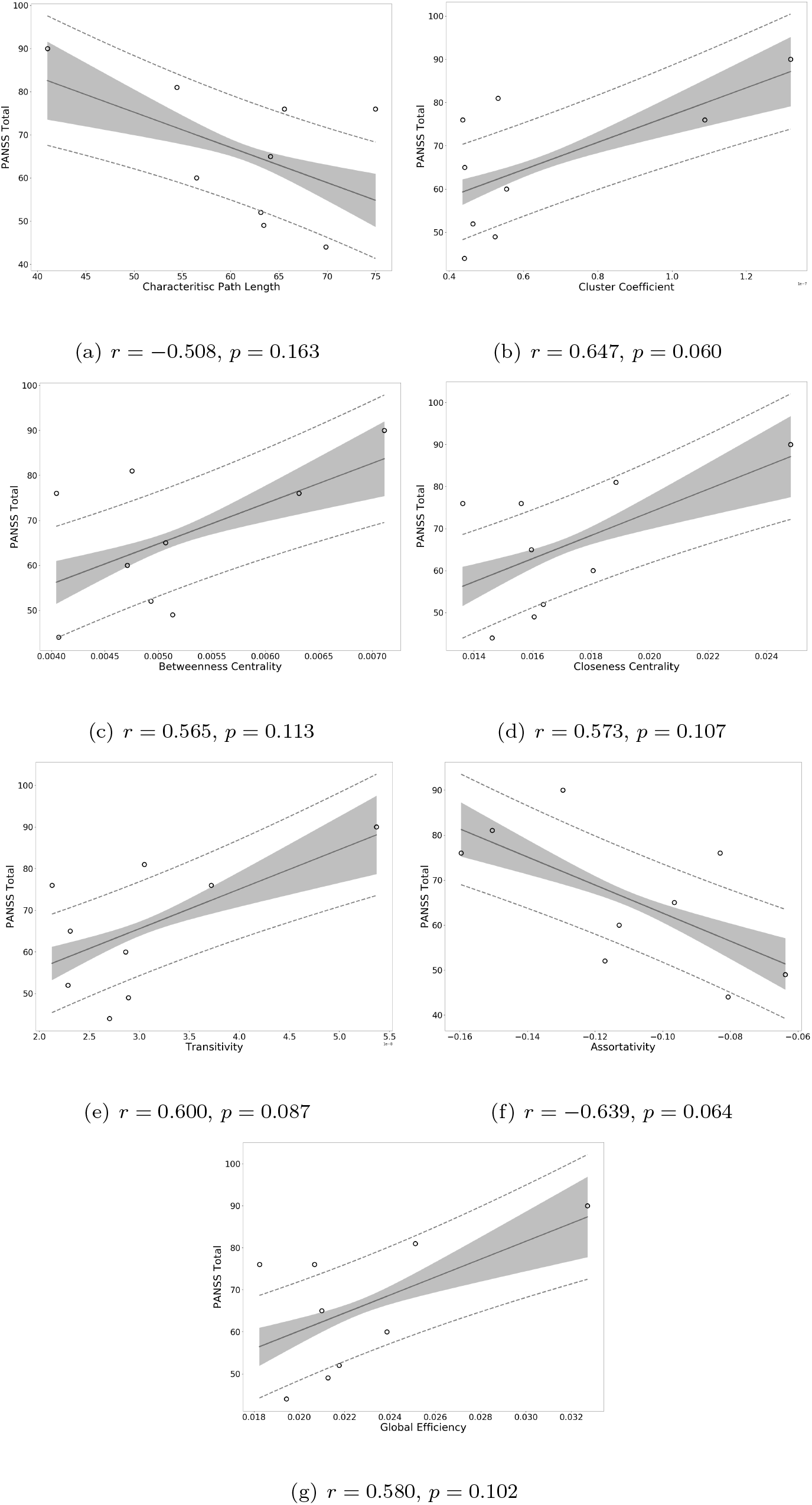
Correlation of graph measures of the SCZaff group with PANSS total scores

**Figure 7:**
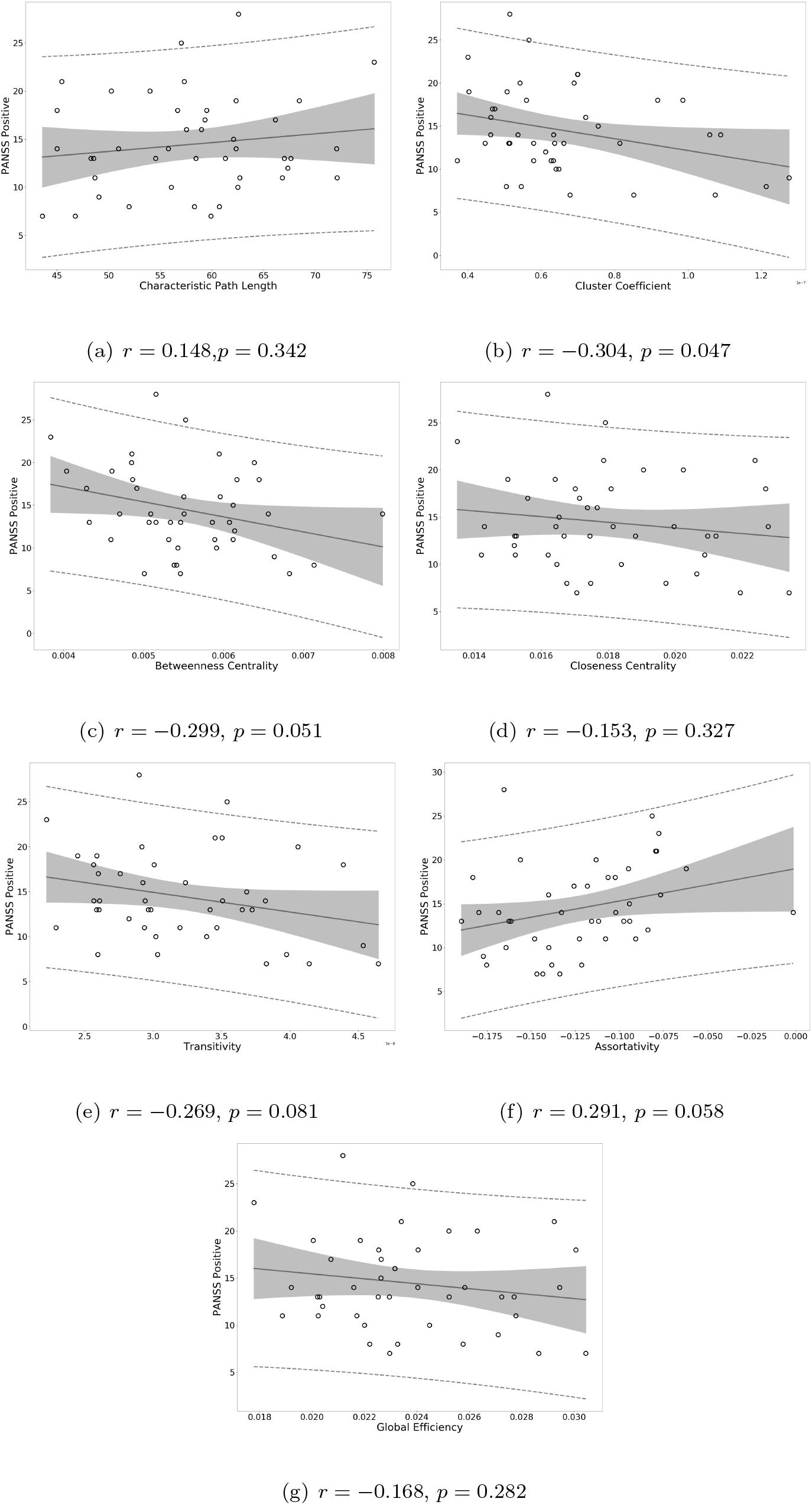
Correlation of graph measures of the SCZ group with PANSS positive scores

**Figure 8:**
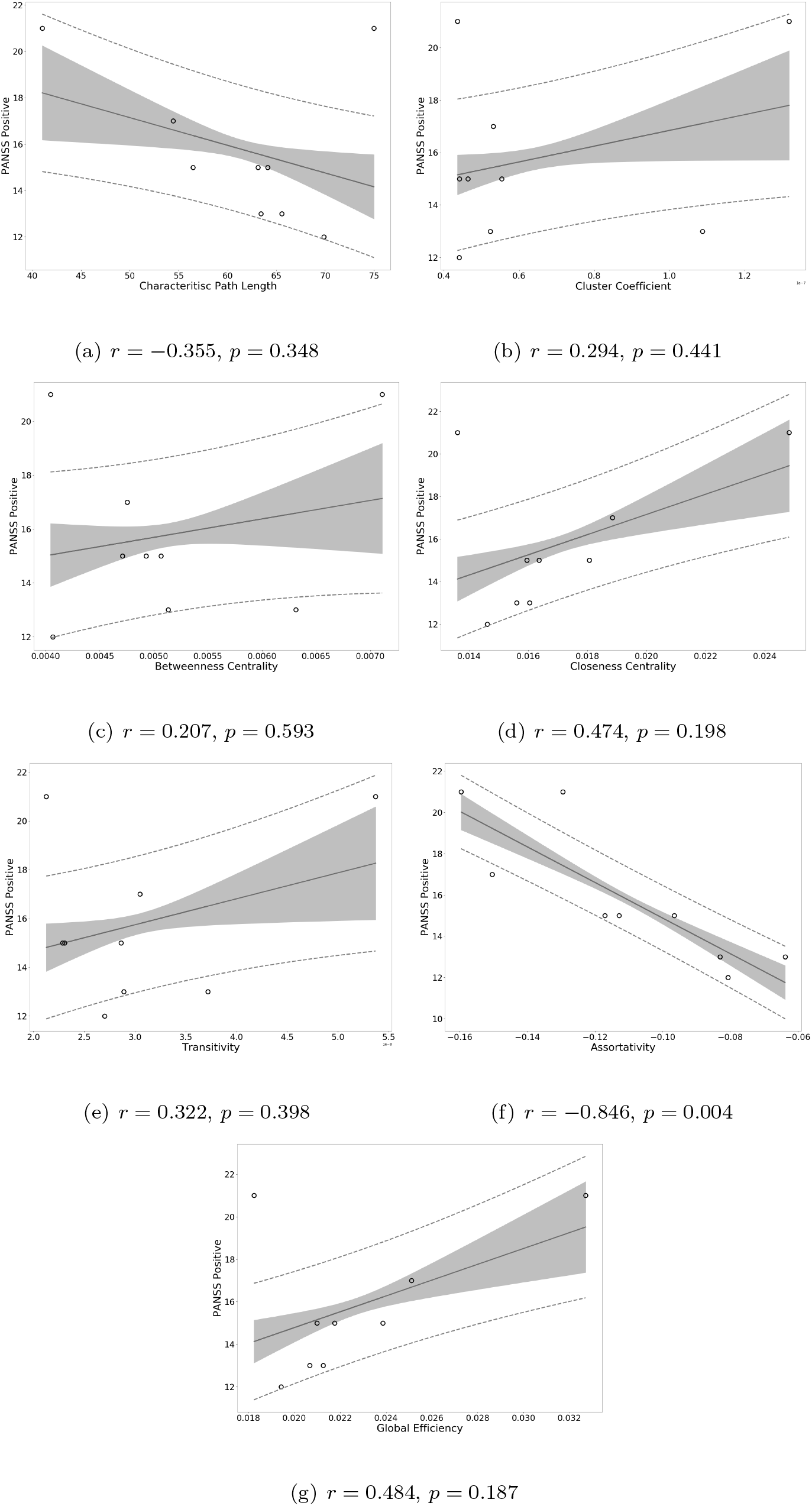
Correlation of graph measures of the SCZaff group with PANSS positive scores.

**Figure 9:**
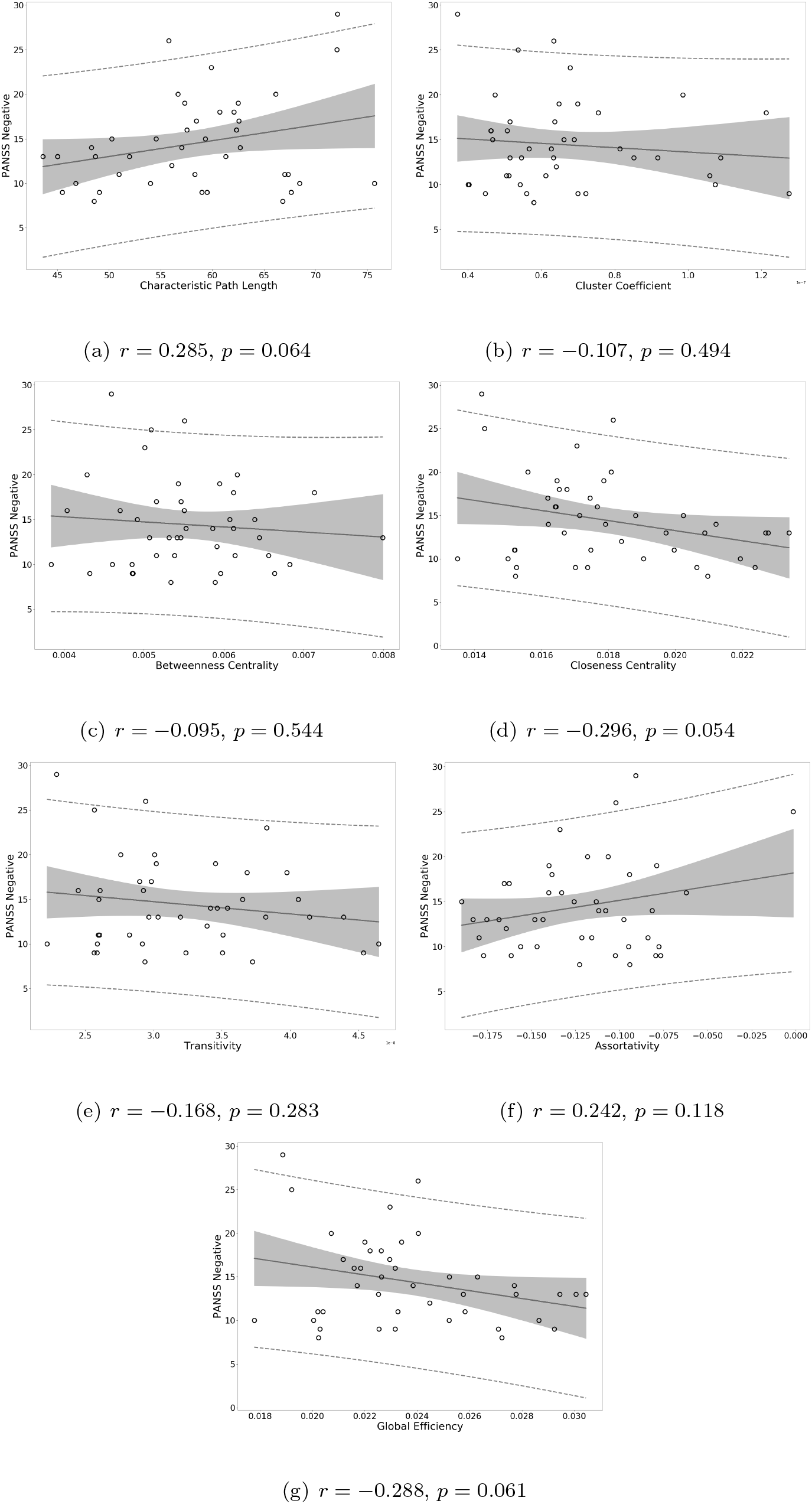
Correlation of graph measures of the SCZ group with PANSS negative scores.

**Figure 10:**
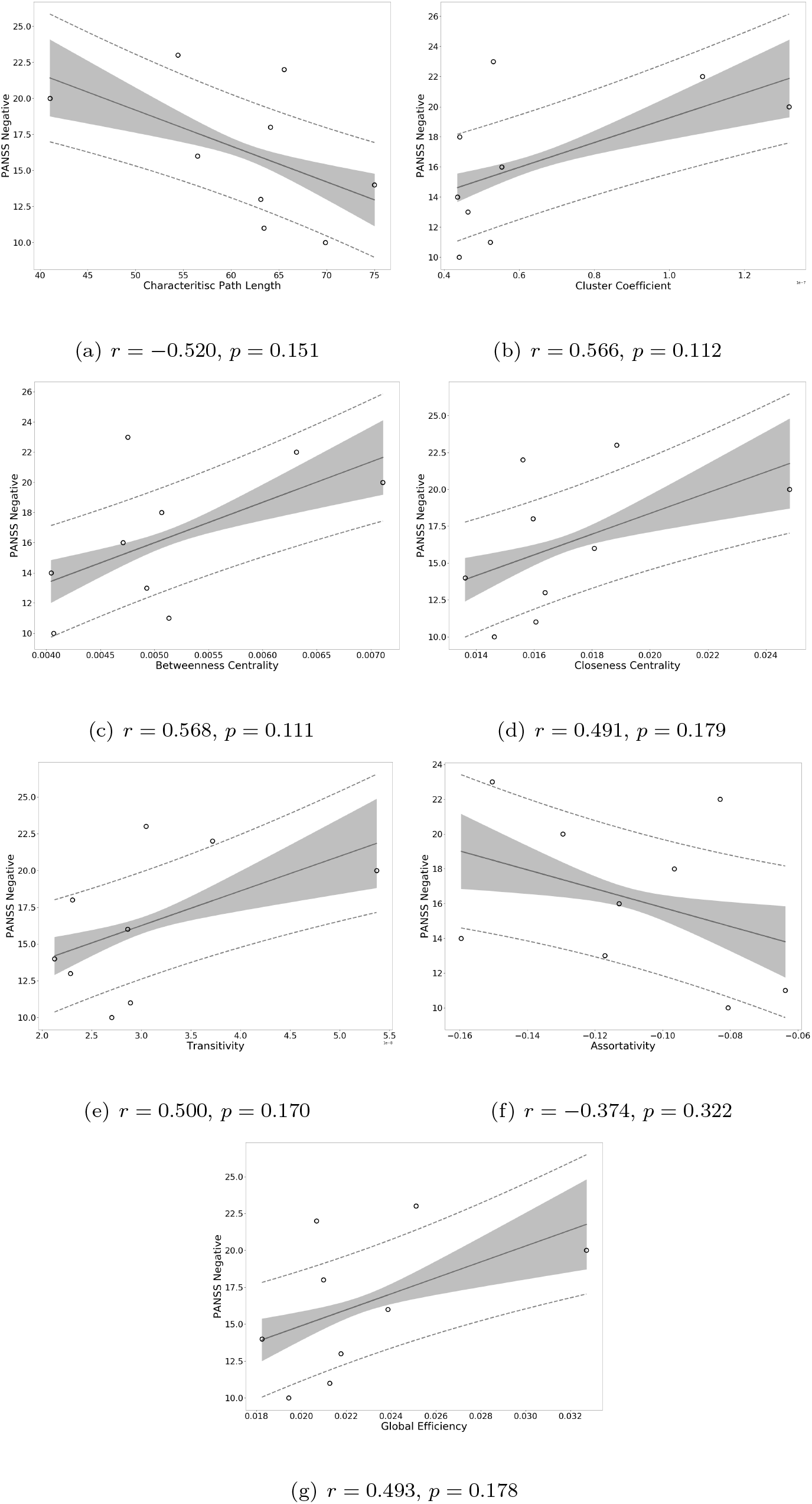
Correlation of graph measures of the SCZaff group with PANSS negative scores.

**Figure 11:**
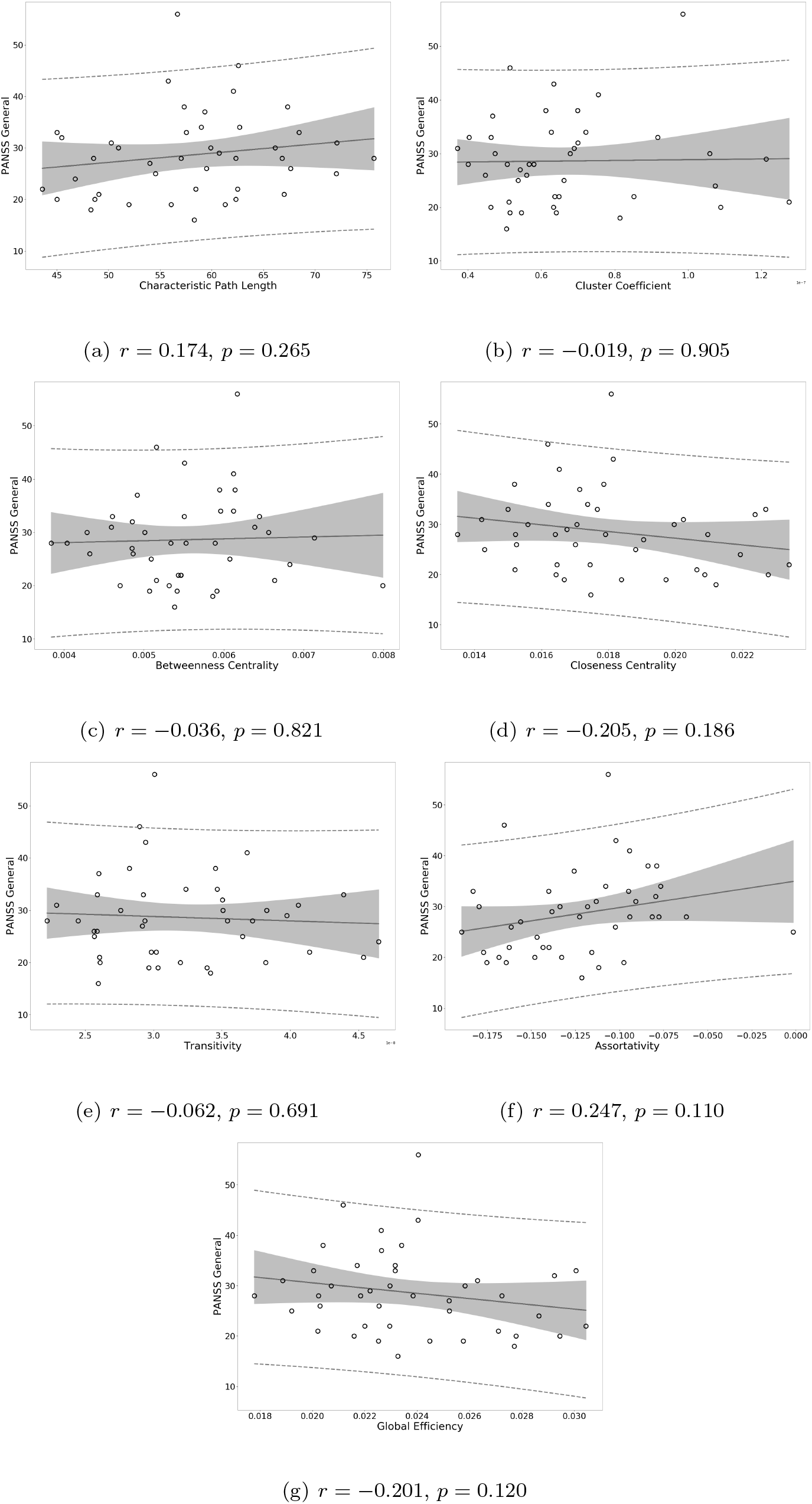
Correlation of graph measures of the SCZ group with PANSS general scores.

**Figure 12:**
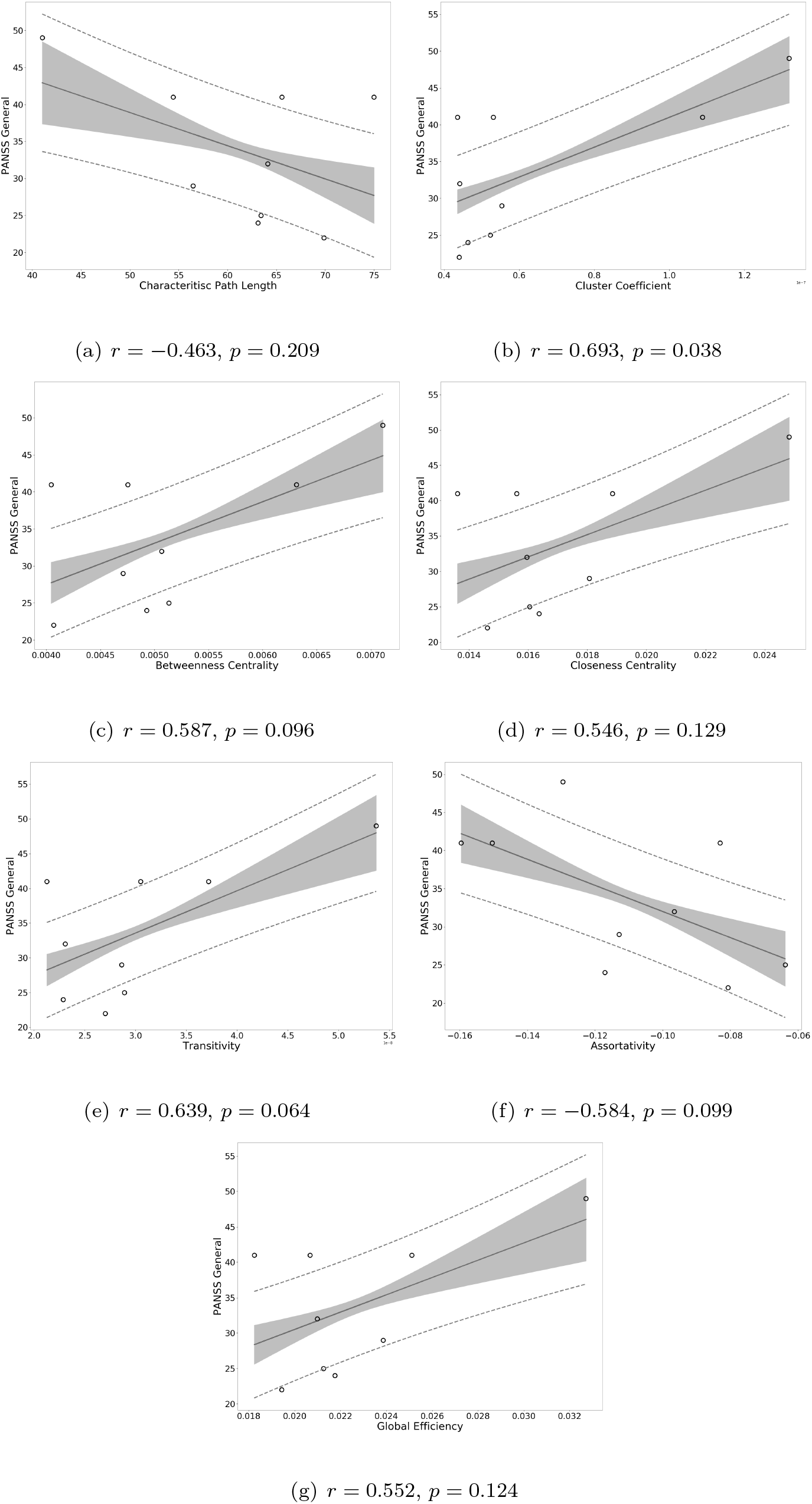
Correlation of graph measures of the SCZaff group with PANSS general scores.

### Control Measures

**Figure 13:**
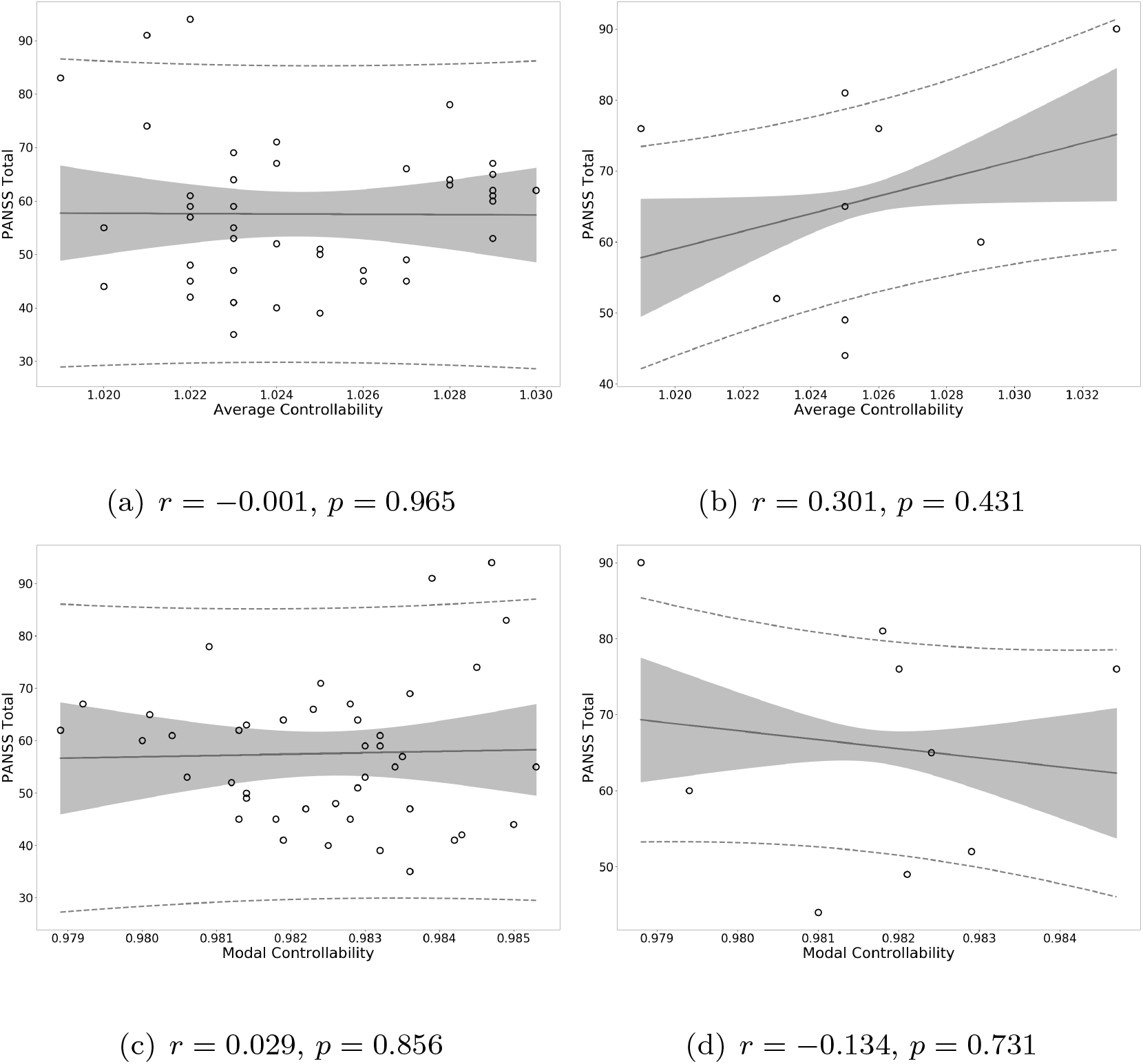
Correlation of controllability measures with PANSS total scores.

**Figure 14:**
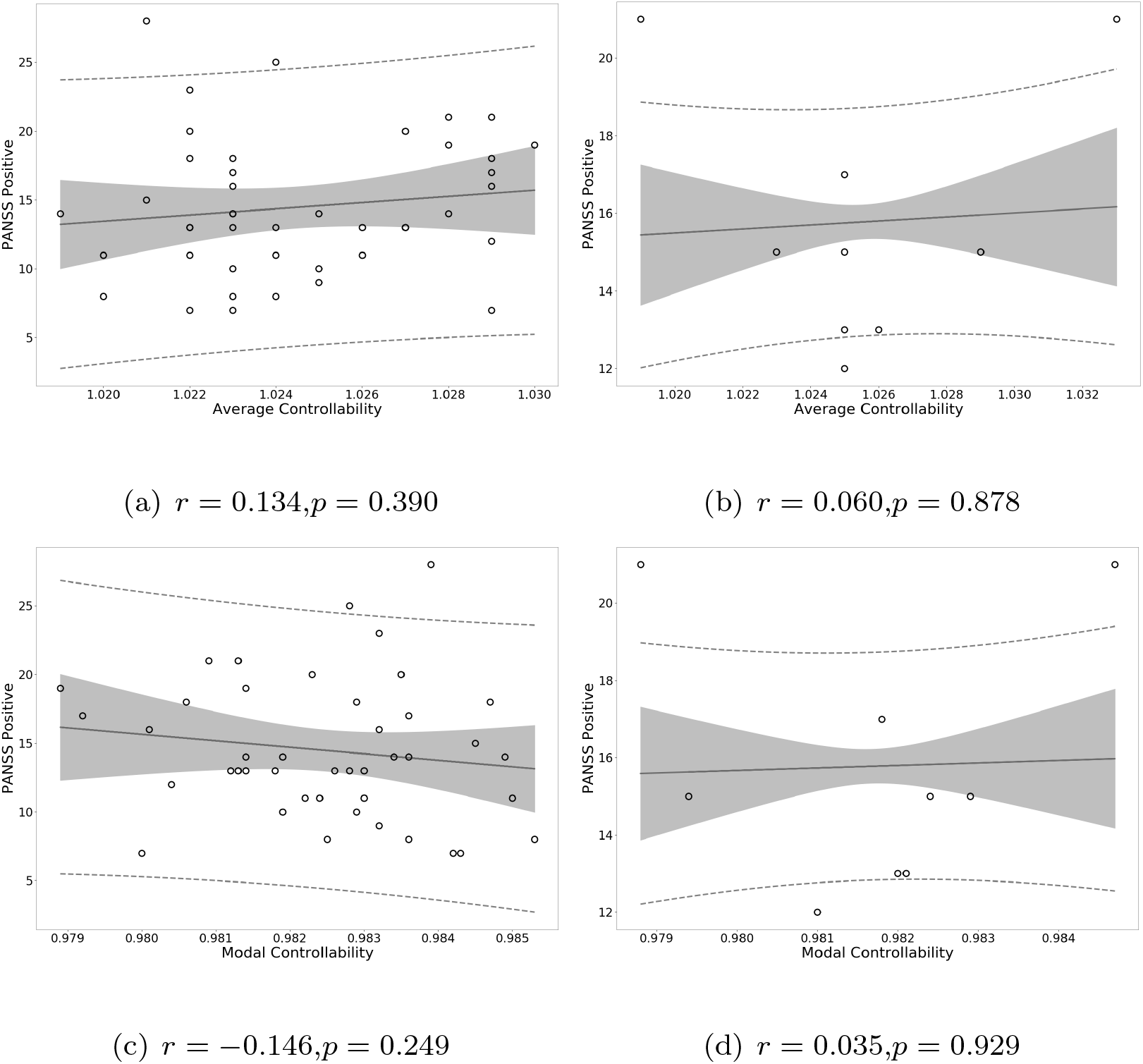
Correlation of controllability measures with PANSS positive scores.

**Figure 15:**
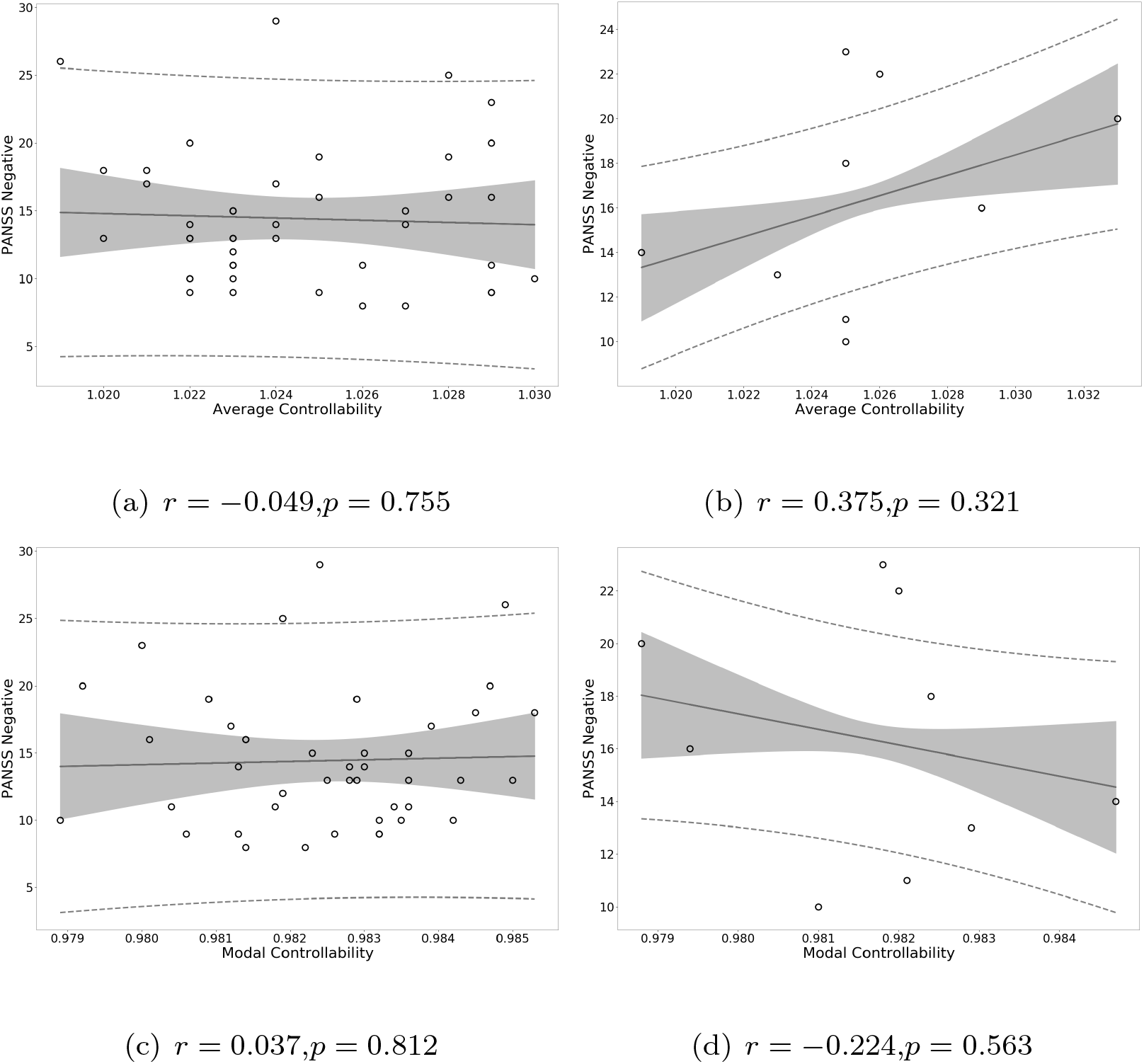
Correlation of controllability measures with PANSS negative scores.

**Figure 16:**
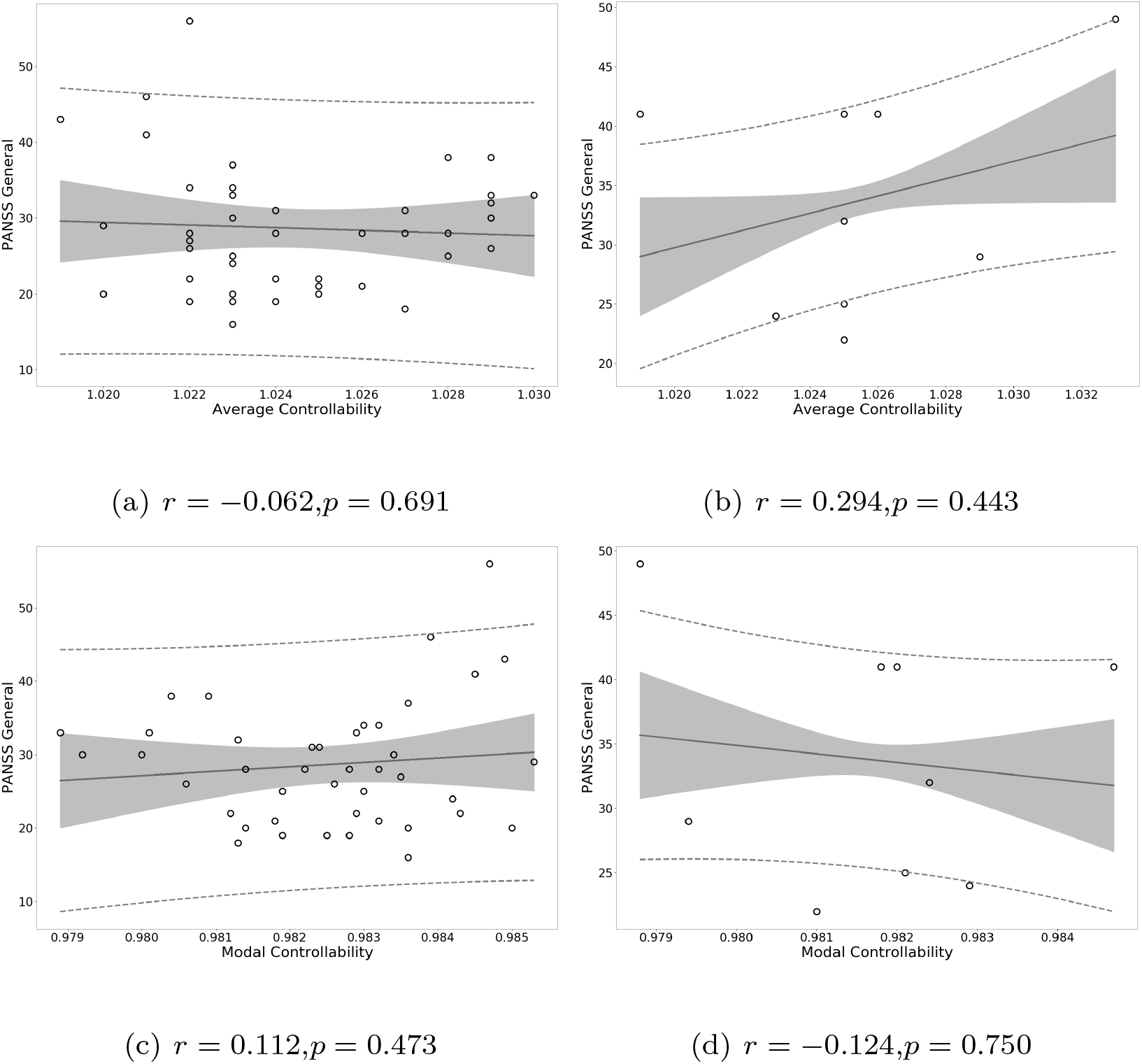
Correlation of controllability measures with PANSS general scores.

**Figure 17:**
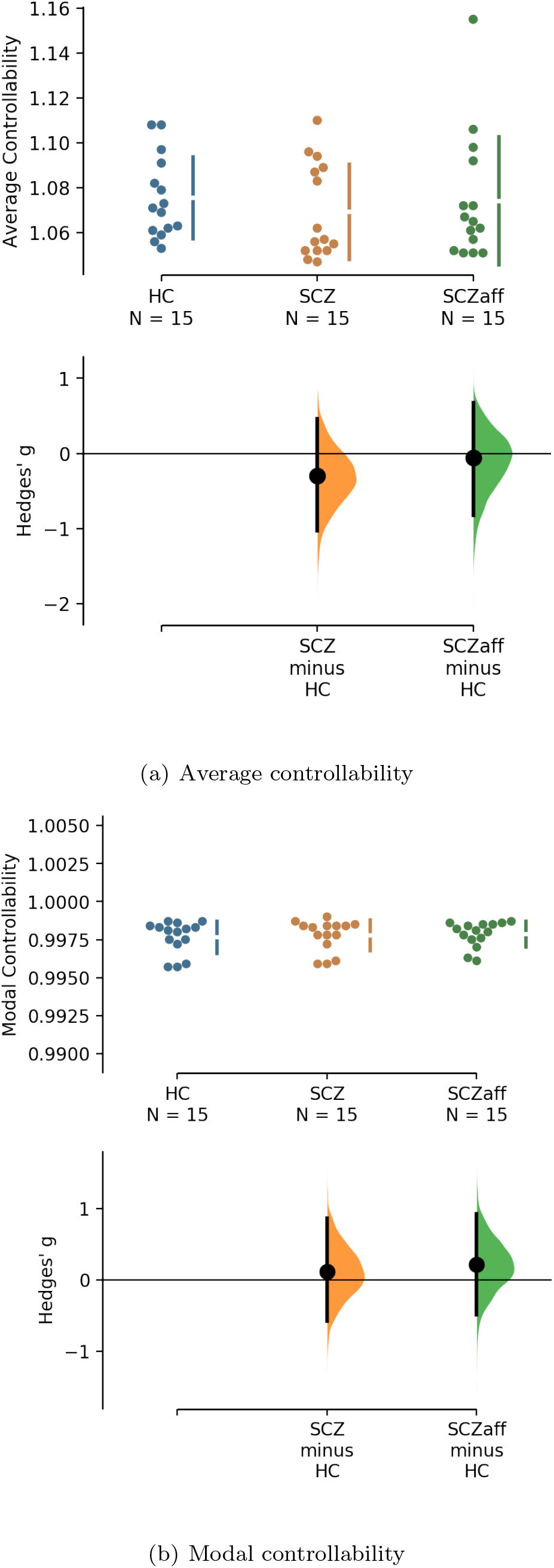
Cognitive systems and controllability

**Table 7:** Table 6, continued. Assignment of AAL2 regions to one of cognitive systems continued: Somatomotor, Default Mode (DMN), Control, Visual, Dorsal Attention, Ventral Attention, Limbic, and Other.

| Region | Cognitive System |
| --- | --- |
| Rectus_L | Limbic |
| Rectus_R | Limbic |
| OFCmed_L | Limbic |
| OFCmed_R | Limbic |
| OFCant_L | Limbic |
| OFCant_R | Control |
| OFCpost_L | Limbic |
| OFCpost_R | Limbic |
| OFClat_L | Default Mode Network |
| OFClat_R | Default Mode Network |
| Insula_L | Ventral Attention |
| Insula_R | Ventral Attention |
| Cingulate_Ant_L | Default Mode Network |
| Cingulate_Ant_R | Default Mode Network |
| Cingulate_Mid_L | Ventral Attention |
| Cingulate_Mid_R | Ventral Attention |
| Cingulate_Post_L | Default Mode Network |
| Cingulate_Post_R | Default Mode Network |
| Hippocampus_L | Other |
| Hippocampus_R | Limbic |
| ParaHippocampal_L | Limbic |
| ParaHippocampal_R | Limbic |

**Table 8:**
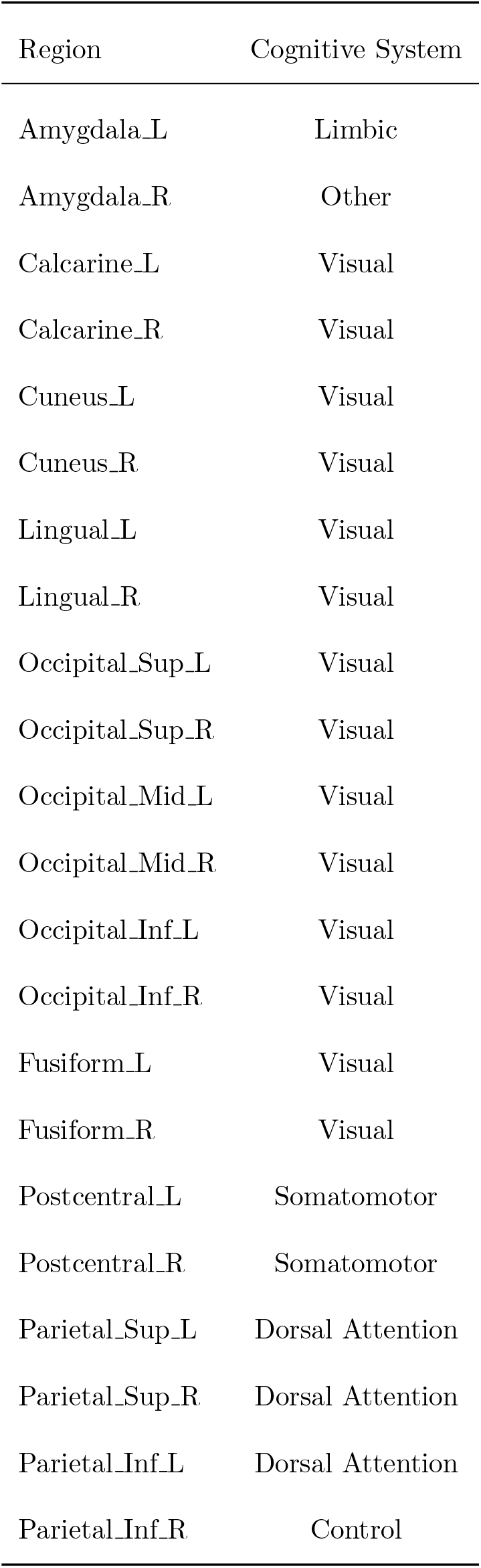
Table 6, continued. Assignment of AAL2 regions to one of cognitive systems continued: Somatomotor, Default Mode (DMN), Control, Visual, Dorsal Attention, Ventral Attention, Limbic, and Other.

**Table 9:** **Table 6, continued**. Assignment of AAL2 regions to one of cognitive systems continued: Somatomotor, Default Mode (DMN), Control, Visual, Dorsal Attention, Ventral Attention, Limbic, and Other.

| Region | Cognitive System |
| --- | --- |
| SupraMarginal_L | Ventral Attention |
| SupraMarginal_R | Ventral Attention |
| Angular_L | Default Mode Network |
| Angular_R | Default Mode Network |
| Precuneus_L | Default Mode Network |
| Precuneus_R | Dorsal Attention |
| Paracentral_Lobule_L | Somatomotor |
| Paracentral_Lobule_R | Somatomotor |
| Caudate_L | Limbic |
| Caudate_R | Other |
| Putamen_L | Other |
| Putamen_R | Ventral Attention |
| Pallidum_L | Other |
| Pallidum_R | Other |
| Thalamus_L | Other |
| Thalamus_R | Other |
| Heschl_L | Somatomotor |
| Heschl_R | Somatomotor |
| Temporal_Sup_L | Somatomotor |
| Temporal_Sup_R | Somatomotor |

**Table 10:**
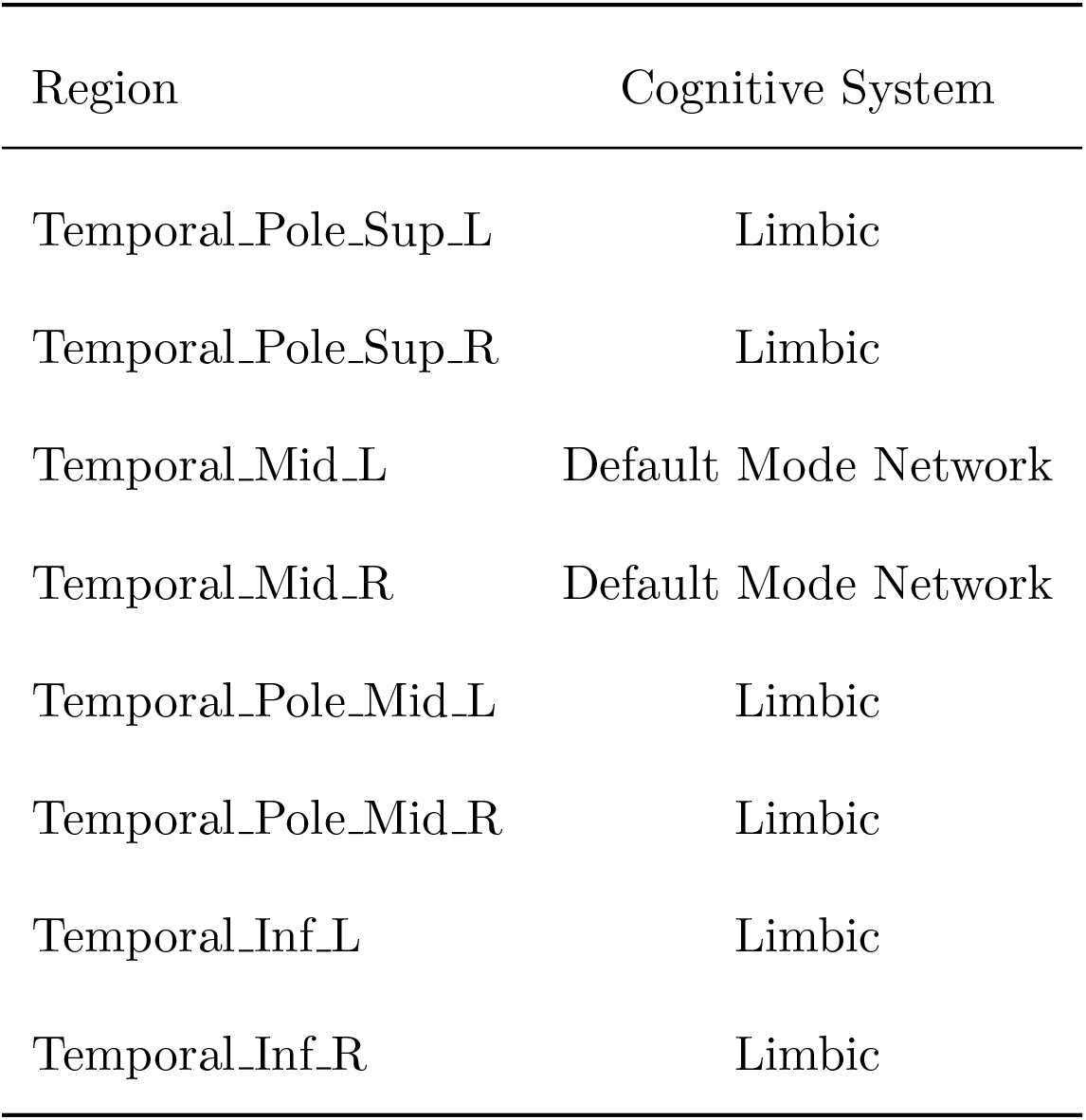
**Table 6, continued**. Assignment of AAL2 regions to one of 8 cognitive systems continued: Somatomotor, Default Mode (DMN), Control, Visual, Dorsal Attention, Ventral Attention, Limbic, and Other.

| Region | Cognitive System |
| --- | --- |
| Temporal_Pole_Sup_L | Limbic |
| Temporal_Pole_Sup_R | Limbic |
| Temporal_Mid_L | Default Mode Network |
| Temporal_Mid_R | Default Mode Network |
| Temporal_Pole_Mid_L | Limbic |
| Temporal_Pole_Mid_R | Limbic |
| Temporal_Inf_L | Limbic |
| Temporal_Inf_R | Limbic |

